# Nationwide melanoma registry databases in real-world settings: a scoping review protocol

**DOI:** 10.1101/2024.08.28.24311481

**Authors:** Songchun Yang, Dilan Deng, Wenrui Lin, Xiaozhen Chen, Shuang Zhao, Lixia Lu, Yi Xiao, Minxue Shen, Mingliang Chen, Xiang Chen, Juan Su

## Abstract

**Introduction:** A patient registry database is an important tool to address a wide range of research questions. Several countries have established nationwide melanoma registry databases. However, there is no report on summarising and comparing these databases. This scoping review aims to answer a broad question on how contemporary nationwide melanoma registry databases were conducted across different countries.

**Methods:** The proposed scoping review will follow the guidelines described by the Joanna Briggs Institute manual for evidence synthesis and results will be reported according to the Preferred Reporting Items for Systematic Review and Meta-Analysis extension for Scoping Reviews guideline. PubMed, Embase, Web of Science, ClinicalTrials.gov, and Google will be parallelly and independently searched by two reviewers. We will include nationwide melanoma databases with the aim of continuously and systematically collecting medical records and other relevant information about melanoma patients under routine conditions. The reference list of all included publications will also be screened for additional literature. The literature search will be limited to records after the year 2000 and in English. Two reviewers will independently review the selected literature for information extraction according to a predefined extraction guidance sheet. For databases without relevant literature introductions, we will try contacting the main researchers of the database via email for detailed information. We will provide a brief descriptive analysis of the collected data.

**Expected results and implications:** This scoping review will provide an extensive description of how contemporary nationwide melanoma databases were conducted. Findings have the potential to enhance the understanding of melanoma registry databases among researchers in related fields and promote subsequent international collaborations that aim at promoting the clinical practice of melanoma.

**STRENGTHS AND LIMITATIONS OF THIS STUDY:**

➢ This is the first scoping review protocol of summarizing contemporary nationwide melanoma databases globally.
➢ Targeted online surveys through email enable us to have a more comprehensive understanding of the latest situation of current databases.
➢ We expect to answer a broad question on how contemporary nationwide melanoma registry databases were conducted across different countries.
➢ Potentially missing relevant databases that have not been insufficiently reported is a limitation of the present scoping review.

## INTRODUCTION

Melanoma is a malignant tumour arising from melanocytes. Cutaneous melanoma (including acral melanoma) accounts for the majority of melanoma diagnoses (>90%), with melanomas of mucosal and uveal origin occurring more rarely.^1^ Globally, it is estimated there were 331,647 new cases of cutaneous melanoma and 58,645 deaths caused by cutaneous melanoma in 2022.^2^ Patient registries are prospective and systematic data collections, which are non-interventional (observational) in nature and reflect care practices under routine conditions (i.e., the real-world settings).^3^ A patient registry database might serve as an important tool to address a wide range of research questions, such as evaluating and comparing the clinical effects of different treatments, exploring factors that might affect the prognosis, developing and validating prognostic models and so on. Also, the patients registered in the database can be identified as potential study participants for randomized controlled trials. The real-world evidence generated by these studies has the potential to guide future clinical practice.

In recent years, some studies have reported on the development of melanoma registry databases in countries such as Spain, Australia, Denmark, and the Netherlands.^4-10^ We have performed a preliminary review of published manuscripts or protocols. We have not found a scoping review or knowledge synthesis on contemporary nationwide melanoma registry databases. Therefore, the objective of this scoping review is to answer a broad question on how contemporary nationwide melanoma registry databases were conducted across different countries.

## METHODS AND ANALYSIS

The proposed scoping review will follow the guidelines described by the Joanna Briggs Institute (JBI) manual for evidence synthesis.^11^ Results will be reported according to the Preferred Reporting Items for Systematic review and Meta-Analysis extension for Scoping Reviews (PRISMA-ScR) guideline.^12^

### Aims and objectives

The overall objective of this scoping review is to identify and summarize contemporary nationwide melanoma registry databases around the world. The current scoping review will be conducted according to the research plan displayed in ***Figure 1***. We search for related literature about melanoma registry databases and extract relevant items from them. When encountering insufficient reports, we will try contacting the person in charge of the database or the corresponding author of the relevant publication and inviting them to supplement the information.

**Figure 1.**
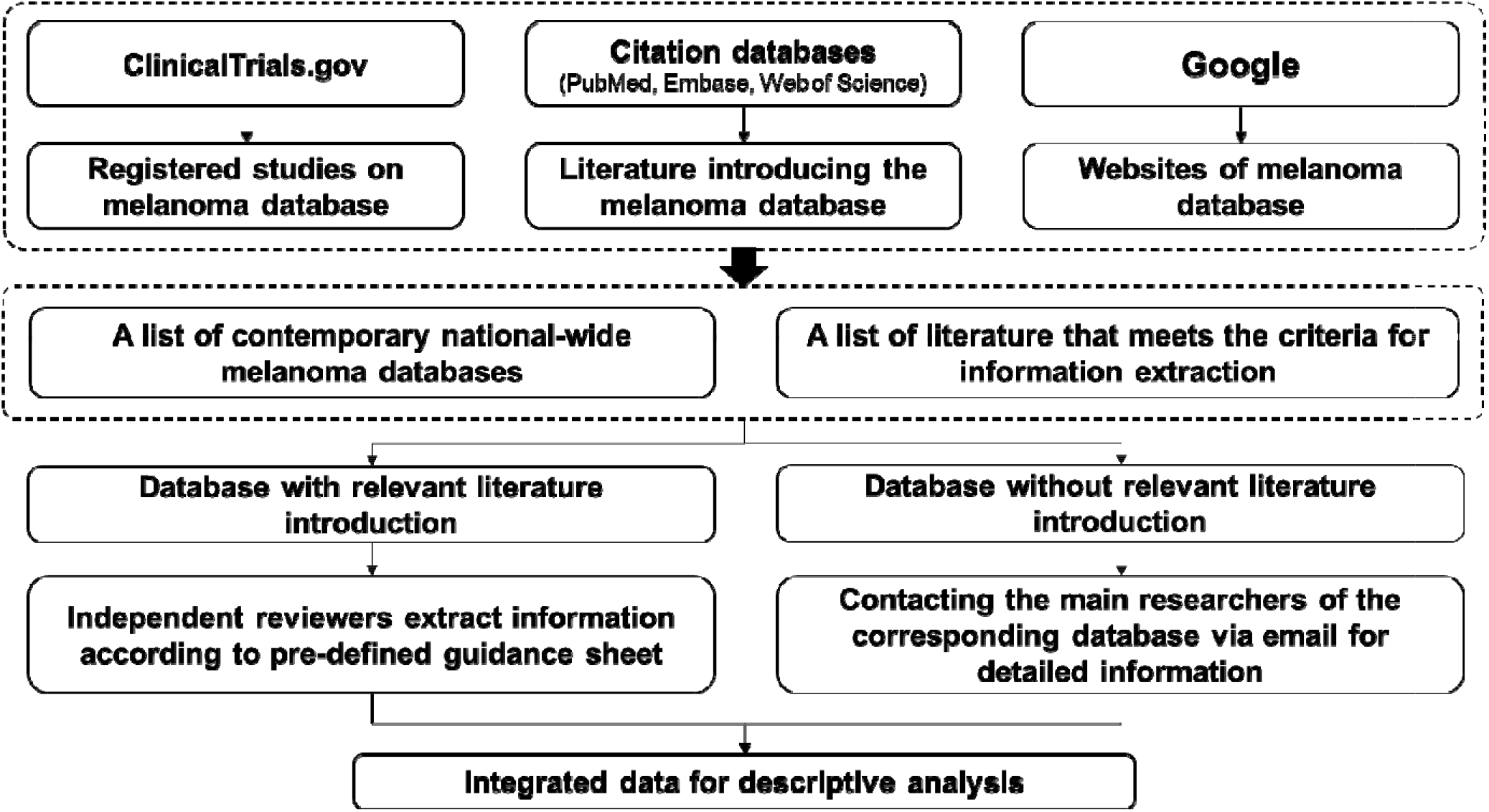
The research plan for the scoping review.

### Patient and public involvement

The present scoping review focuses on the database of melanoma patients, not melanoma patients themselves. Therefore, no patients or the public were recruited for this protocol. Patients or the public were not involved in the design, development or conduct of this current research protocol. We will introduce the main findings of this work through the internet after the publication of this scoping review. This knowledge might be useful to guide the building of subsequent national or regional melanoma registries.

### Eligibility criteria

In the present scoping review, we will include databases that meet the following criteria: (1) it should be a nationwide database of melanoma, and (2) the aim of establishing the database is to continuously and systematically collect medical records and other relevant information of melanoma patients under routine conditions, usually not limited to a specific research objective. We will exclude the following databases: (1) those are not established specifically for melanoma (such as the National Cancer Registry); (2) those only cover a few regions of a country; and (3) those are temporarily collected to answer one or two research questions and the researchers usually do not have a long-term maintenance plan for it. The melanoma patients included in the database could be at any tumor stage and any melanoma subtypes (i.e., cutaneous, acral, mucosal, uveal, etc.). Other inclusion and exclusion criteria are summarized in ***Table 1***.

**Table 1.** Summary of inclusion and exclusion criteria.

| **Database characteristics** | **Inclusion criteria** | **Exclusion criteria** |
| --- | --- | --- |
| Aim of the database | To continuously and systematically collect medical records and other relevant information (such as questionnaires, prognosis, adverse reactions, bio-samples, etc.) of melanoma patients. Usually not limited to a specific research objective. | ● Databases that are not established specifically for melanoma, such as the National Cancer Registry.<br>● Databases that temporarily collected to answer one or two specific research questions and the researchers usually do not have a long-term maintenance plan for it. |
| Patients included in the database | ● Melanoma of any origin (skin, mucosal, uveal, etc.)<br>● Melanoma at any tumor stage | None |
| Age limits of patients in the database | All ages | None |
| Study designs | Cohort study | ● Case-control study<br>● Cross-sectional study<br>● Randomized clinical trial |
| Publication types | All publication types | Grey literature |
| Publication status | Published or preprint | None |
| Countries | All countries | None |
| Dates | All literature from Jan 1, 2000 onwards | Literature before Jan 1, 2000 |
| Languages | English | Non-English |

### Information sources

In the present scoping review, we will search the melanoma databases through multiple channels, which include: (1) PubMed (pubmed.ncbi.nlm.nih.gov), Embase (www.embase.com), and Web of Science (webofscience.clarivate.cn/wos); (2) the Clinical Trials database (clinicaltrials.gov); and (3) Google (www.google.com). Two authors conducted searches on various platforms based on predetermined search terms. The reference list of all included publications will also be screened for additional literature. Studies that are registered in the Clinical Trial database will be searched to identify the melanoma databases that are in the initial stage of establishment. When using the search engine Google for retrieval, the main purpose is to identify the melanoma database that has not been formally reported in any published papers but has established a dedicated website. The pilot searches in PubMed were started on July 28, 2024. Details of the initial search in PubMed is provided in the ***Table 2***. Formal searches in all the selected databases will be run on September 15, 2024.

**Table 2.**
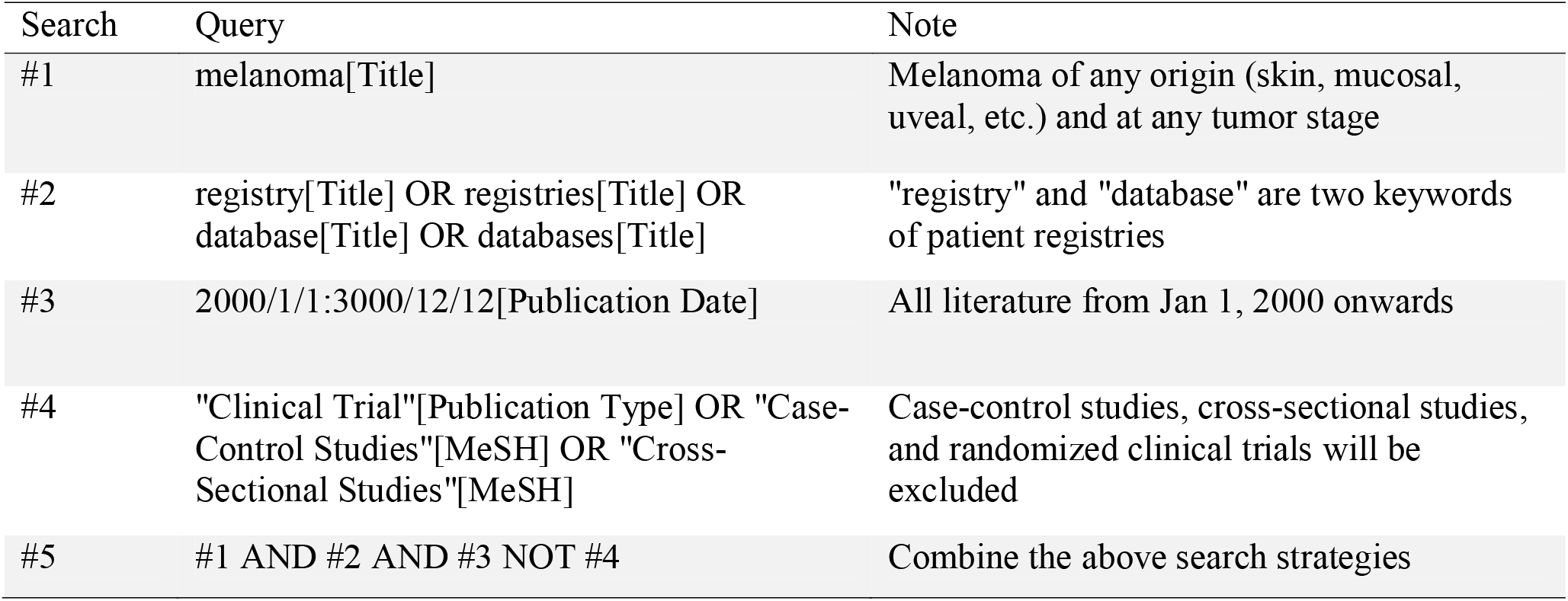
Search strategy terms for PubMed.

| Search | Query | Note |
| --- | --- | --- |
| #1 | melanoma[Title] | Melanoma of any origin (skin, mucosal, uveal, etc.) and at any tumor stage |
| #2 | registry[Title] OR registries[Title] OR database[Title] OR databases[Title] | "registry" and "database" are two keywords of patient registries |
| #3 | 2000/1/1:3000/12/12[Publication Date] | All literature from Jan 1, 2000 onwards |
| #4 | "Clinical Trial"[Publication Type] OR "Case-Control Studies"[MeSH] OR "Cross-Sectional Studies"[MeSH] | Case-control studies, cross-sectional studies, and randomized clinical trials will be excluded |
| #5 | #1 AND #2 AND #3 NOT #4 | Combine the above search strategies |

### Selection process

All searched literature will be collated and imported into EndNote software. The duplicates will be removed. The literature search will be limited to records after the year 2000 and in English. Two authors will review the retrieved titles and abstracts independently to identify literature that may contain information on nationwide melanoma databases. The selected papers will be further examined to see if they meet the conditions for formal information extraction. Any inconsistencies that arise during the inclusion and exclusion of literature will be resolved after discussion. The final process of including and excluding literature will be documented and reported in the form of a flowchart in the scoping review. In the end, we will obtain (1) the names of all retrieved melanoma databases and (2) a list of literature that meets the criteria for information extraction.

### Data collection process

We have designed a data extraction guidance to clarify the information that needs to be extracted from the literature (***appendix 1***). For each retrieved melanoma database, if there is literature specifically introducing the database, two authors will independently review the related literature for information extraction according to the predefined extraction guidance sheet. For databases without relevant literature introductions or with insufficient reporting, we will try contacting the main researchers of the database via email and inviting them to fill out an online survey form regarding the database (https://www.wjx.cn/vm/h5m7Xq1.aspx). Formal email invitations are planned to be sent out from October 1, 2024, to November 30, 2024. The data obtained through two methods will ultimately be merged into one database.

### Statistical analysis

We will provide a brief descriptive analysis of the collected data, including the median sample size of different melanoma databases, the proportion of databases containing acral melanoma and mucosal melanoma, the proportion of databases that have collected biological samples, the proportion of databases that are still collecting data prospectively, etc. We will use appropriate figures or charts to display the results. All missing values will be reported.

### Quality appraisal

In the present scoping review, we will not conduct a quality appraisal. This is because the objective of this scoping review is to identify all contemporary nationwide melanoma databases and summarize their features other than to evaluate the quality of data collected in these databases or to evaluate the report quality of the literature introducing these databases.

### Strengths and Limitations

To the best of our knowledge, this is the first scoping review protocol for summarizing contemporary nationwide melanoma databases. The combination of direct information extraction from the published literature and targeted online surveys through email is expected to enable us to have a more comprehensive understanding of the latest situation of current databases. Potentially missing relevant databases that have not been insufficiently reported is a limitation of the present scoping review. We will search the melanoma databases through multiple channels to minimize such omissions. In addition, email surveys might not have an ideal response rate, which will limit our ability to obtain more detailed data.

### Ethics and dissemination

This study does not directly involve any human participants as it concentrates on the database itself. Therefore, ethics committee approval is not required for this study.

The findings of our review will aim to be published in a peer-reviewed journal.

## CONCLUSION

The objective of this scoping review is to identify and summarize contemporary nationwide melanoma databases around the world. This review has the potential to enhance the understanding of melanoma registry databases among researchers in related fields and promote subsequent international collaborations that aim at promoting the clinical practice of melanoma.

## Data Availability

All data produced in the present study are available upon reasonable request to the authors

## Contributors

XC and JS are joint senior authors and designed the study. SY wrote the first draft of the manuscript. SY and DD conducted search strategy development. JS and XC supervised and obtained funding for the study. JS, XC, and SY had full access to all the data in the study and took responsibility for the integrity of the data and the accuracy of the data analysis. JS is the guarantor. JS, XC, MC, MS, YX, LL, SZ, XZC, WL, and DD critically revised the manuscript. The corresponding author attests that all listed authors meet authorship criteria and that no others meeting the criteria have been omitted.

## Patient and public involvement

Patients and/or the public were not involved in the design, conduct, reporting, or dissemination plans of this research.

## Data availability statement

Further information is available from the corresponding author upon request.

