## Appendix 1 for "Nationwide melanoma registry databases in real-world settings: a scoping review protocol"

**Appendix 1. Data Extraction Guidance**

**To cite:** Yang S, Deng D, Lin W, *et al*. Nationwide melanoma registry databases in real-world settings: a scoping review protocol. *medRxiv* 2024. doi:

* Required

1. Country *

_________________________________

2. Full name of the melanoma database *

Please use the name **in English**

_________________________________

3. Abbreviation name of the melanoma database

_________________________________

4. Year when the database was officially set up *

Please note that this **does not** refer to the time of the earliest patient in the database.

_________________________________

5. Does the database include confirmed melanoma cases before establishment? *

*Mark only one oval*.

Some databases **retrospectively** include historical medical records of melanoma patients **before they are established**. Does the current database have this situation?

○Yes

○No

6. Has the current database participated in any international collaboration groups (such as the European Melanoma Registry)? *

*Mark only one oval*.

○Yes

○No

7. (**If the answer of question 6 is "Yes"**) What are the names of these international collaboration groups?

_________________________________

8. What is the sample size of the latest version of the database? *

If it's not precise, you can fill in the approximate number of melanoma patients.

_________________________________

9. Is the above sample size an exact number or a rough number? *

*Mark only one oval*.

○Exact number

○Rough number

10. The release date of the latest version of the database *

_________________________________

11. Is the database still collecting data prospectively at present? *

*Mark only one oval*.

○Yes

○No

12. (**If the answer of question 11 is "No"**) Time when the database stopped prospectively collecting data

_________________________________

13. The number of hospitals or institutions that are responsible for collecting data *

*Mark only one oval*.

○1~5

○6~10

○11~15

○16~20

○＞20

14. Does the database have a website to introduce the basic information and progress of it? *

*Mark only one oval*.

○Yes

○No

15. (**If the answer of question 14 is "Yes"**) The website of the database

_________________________________

16. The melanoma staging system that is currently used in the database *

*Mark only one oval*.

AJCC: American Joint Committee on Cancer

○AJCC 7th edition

○AJCC 8th edition

○Other (please specify) _________________ *

17. **Tumor stage at recruitment** - The current database includes the following melanoma patients: *

*Multiple options can be selected.*

□In situ

□Stage I

□Stage II

□Stage III

□Stage IV

18. **Tumor subtypes at recruitment** - The current database includes the following melanoma patients: *

*Multiple options can be selected.*

□Cutaneous melanoma (excluding acral melanoma)

□Acral melanoma

□Mucosal melanoma

□Uveal melanoma

□Other subtypes

19. Is there a minimum age limit when including patients in the current database? *

*Mark only one oval*.

○Yes (please specify below)

○No

20. (**If the answer of question 19 is "Yes"**) The minimum age limit

_________________________________

21. Is there a maximum age limit when including patients in the current database? *

*Mark only one oval*.

○Yes (please specify below)

○No

22. (**If the answer of question 21 is "Yes"**) The maximum age limit

_________________________________

23. Are there any other inclusion criteria when including patients in the current database? *

*Mark only one oval*.

○Yes (please specify below)

○No

24. (**If the answer of question 23 is "Yes"**) Other inclusion criteria *

Please describe other inclusion criteria here

_________________________________

25. Are there any **exclusion criteria** when including patients in the current database? *

*Mark only one oval*.

○Yes (please specify below)

○No

26. (**If the answer of question 25 is "Yes"**) The exclusion criteria

Please describe the exclusion criteria here

_________________________________

27. Are there any published papers or documents specifically introducing the database? *

*Mark only one oval*.

○Yes (please specify below)

○No

28. (**If the answer of question 27 is "Yes"**) How to access these papers or documents?

*PMID or DOI*

_________________________________

29. Which of the following data are available in the current database? *

*Mark only one oval* *for each row.*

| Items | Available | Not available | Not sure |
| --- | --- | --- | --- |
| Occupation | ○ | ○ | ○ |
| Education | ○ | ○ | ○ |
| Household income | ○ | ○ | ○ |
| Physical examination of the skin lesions (location, size, color, etc.) | ○ | ○ | ○ |
| Detailed information on pathological examination | ○ | ○ | ○ |
| Detailed information on laboratory tests | ○ | ○ | ○ |
| Detailed information on imaging examination | ○ | ○ | ○ |
| Detailed information on treatments | ○ | ○ | ○ |
| Utilization of outpatient and inpatient medical services before diagnosis | ○ | ○ | ○ |
| Patient's lifestyle (smoking, alcohol drinking, physical activity, diet, etc.) | ○ | ○ | ○ |
| Patient's history of other major chronic diseases (coronary heart disease, stroke, diabetes, etc.) | ○ | ○ | ○ |
| Patient's emotional state (anxiety, depression) | ○ | ○ | ○ |
| Tumor progression and remission during follow-up | ○ | ○ | ○ |
| Adverse reactions after treatment during follow-up | ○ | ○ | ○ |
| Death events during follow-up | ○ | ○ | ○ |

30. Have any biological samples been collected and stored at baseline or during follow-up? *

*Mark only one oval* *for each row.*

| Items | Only at baseline | Only during follow-up | Both | None |
| --- | --- | --- | --- | --- |
| Plasma/serum | ○ | ○ | ○ | ○ |
| Blood cells isolated from whole blood | ○ | ○ | ○ | ○ |
| Urine | ○ | ○ | ○ | ○ |
| Feces | ○ | ○ | ○ | ○ |
| Surgically resected melanoma tissue | ○ | ○ | ○ | ○ |

31. The core research institutions currently responsible for managing and maintaining the database *

_________________________________

32. Does the current database welcome collaboration with research teams from other countries? *

*Mark only one oval*.

○Yes

○No

○Not sure
